# Association of Muscle Strength to Body Composition Measures using DXA, D_3_Cr, and BIA in Collegiate Athletes

**DOI:** 10.1101/2023.05.17.23288849

**Authors:** Devon Cataldi, Jonathan P. Bennett, Brandon K. Quon, Lambert Leong, Thomas L Kelly, William J Evans, Carla M. Prado, Steven B. Heymsfield, John A Shepherd

**Affiliations:** College of Education: Kinesiology and Rehabilitation Science, University of Hawai’i; Department of Epidemiology, University of Hawai’i Cancer Center, 701 Ilalo St, Honolulu, Hawaii, 96813 USA; University of Hawaii Cancer Center, Honolulu, HI, USA; Hologic Inc., Marlborough, MA, USA; Department of Nutrition Sciences, University of California, Berkeley; Department of Agricultural, Life and Environmental Sciences, University of Alberta, Edmonton, AB, Canada; Pennington Biomedical Research Center, Louisiana State University, 6400 Perkins Rd, Baton Rouge, Louisiana, 70808 USA

**Keywords:** D_3_Cr dilution, dual-energy x-ray absorptiometry, body composition, athletic performance, muscle strength, muscle mass

## Abstract

**Background:** Measurements of body composition are helpful indicators of health outcomes, but muscle strength has a greater correlation with disease risk and long-term health outcomes, particularly among older adults. Whole-body DXA scans uniquely parse out total and regional lean soft tissue (LST) and appendicular (ALST), primarily composed of skeletal muscle and often used to diagnose sarcopenia and frailty. An alternative approach measures the enrichment of deuterated Creatinine (D_3_Crn) in urine after ingesting a tracer dose of deuterated creatine (D_3_Cr) to determine creatine pool size and estimate whole-body muscle mass. The utility of D_**3**_Cr relationships between strength and body composition in young athletes has yet to be established. In this study, we investigated the association of muscle strength and body composition using multiple methods including DXA, D_3_Cr, and bioelectrical impedance (BIA), in a collegiate athletic population.

**Methods:** The *Da Kine* Study enrolled 80 multi-sport collegiate athletes. Each subject consumed a 60 mg dose of D_3_Cr and completed whole-body DXA, BIA, and strength tests of the leg and trunk using an isokinetic dynamometer. The analysis was stratified by sex. Pearson’s correlations, forward stepwise linear regression and quartile *p* trend significance were used to show the associations of body composition measures to muscle strength.

**Results:** The mean (SD) age of the 80 (40M/40F) athletes was 21.8 (4.2) years. Raw whole-body values had higher correlations with muscle strength in both sexes compared to the normalized values by height, body mass (BM), and BMI. DXA LST had the highest leg (*R*^*2*^=0.36, 0.37) and trunk (*R*^*2*^=0.53, 0.61) strength in both males and females. Trunk strength was more highly associated with body composition measures than leg strength in both sexes and all measurement techniques. One or more DXA LST measures (total, leg, and ALST) were consistently more highly associated with leg and trunk strengths for both sexes than BIA and D_3_Cr measures. Adjusting all body composition values by age, BMI, and BIA variables did not improve the associations. A significant *p* trend across quartiles was observed for DXA LST and ALST for all measures of strength in both sexes.

**Conclusion:** Although statistical significance was not reached between devices, DXA body composition output variables, especially LST, showed the highest associations with both sexes’ leg and trunk muscle strength. Furthermore, without adjustment for demographic information or BIA variables, whole-body values show stronger associations with muscle strength. Future research should investigate the impact of muscle mass changes on LST and functional measures.

## Introduction

The decline in skeletal muscle mass and strength with age is associated with the functional decline that typically characterizes dynapenia, sarcopenia, and frailty [1]. These structural and functional-related changes in skeletal muscle or appendicular lean soft tissue (ALST) are accompanied by an increased risk of morbidity and mortality [2]. Despite conflicting cut points and adjustments for defining sarcopenia, several definitions rely on measuring muscle mass or its derivatives such as ALST [3].

Many methods exist in determining ALST and/or muscle mass including bioimpedance analysis (BIA), ultrasound, computerized tomography (CT), and magnetic resonance imaging (MRI), but the most commonly utilized method is dual-energy X-ray absorptiometry (DXA) [4]. DXA provides a clinical advantage over CT and MRI due to being broadly available in the US and Europe for osteoporosis assessment [5]. However, DXA has setbacks such as subjecting individuals to low-level radiation and a lack of direct measurement of skeletal muscle. DXA LST includes other tissues that are not made up of pure skeletal muscle, such as tendinous, cartilaginous, and truncal organic tissues and their contents, such as the gut, which can provide a misleading estimate of lean soft tissue (LST). The use of ALST minimizes these errors and hence provides a more accurate estimate of skeletal muscle. ALST by DXA is highly predictive of total body skeletal muscle mass as quantified by MRI (*R*^*2*^=0.96) and CT (*R*^*2*^=0.97) [6].

DXA systems have become progressively more available within elite and professional sports environments, however, DXA does not directly measure muscle mass or strength, but rather through its association with LST, which may explain its limited performance in predicting fall or fracture risk [7]. DXA LST has not been as strongly associated with functional measures, including falls, fractures, mortality, and disability, however, the measure is supposed to be a surrogate of muscle strength, with the combination of DXA ALST and BIA has been suggested to provide insight into both muscle quantity and quality [8]. Muscle quality captures the physiological and metabolic state of muscle tissue and is theorized to explain the loss of muscle function before the loss of muscle mass [9]. There is no agreement on the definition between measures of muscle quality; however, fatty or connective tissue infiltration within muscle tissue has been proposed as measures of muscle quality. [10]. The inclusion of BIA phase angle, as a surrogate measure of muscle quality, appears to improve DXA ALST in its association with grip and lower limb power in both males and females [9, 11] and could potentially provide the same results in athletes. Lastly, DXA LST consists of both protein and water components such that changes in hydration status may obscure true functional muscle mass changes (i.e. the protein) between subjects. For example, it has been observed that regular exercise training, particularly in a warm environment can result in up to 20% plasma volume expansion [12], or other words increasing body hydration.

A novel method for measuring muscle mass, Deuterated D_3_ Creatine (D_3_Cr), has recently been studied against other conventional methods in its accuracy and utility in quantifying muscle mass and strength [13, 14]. A recent review outlines the specific methodologies and assumptions used in the D_3_Cr method for assessing muscle mass [13]. Measuring muscle mass by D_3_Cr has several advantages over DXA including no radiation exposure, cost-effectiveness, and its practicality for both field and at-home settings. However, D_3_Cr can only provide estimates of total body skeletal muscle mass and no other body composition estimates. Furthermore, a considerable amount of time is required for D_3_Cr dilution, sample collection, and analysis (>three days to weeks), to provide a result. Finally, the D_3_Cr dilution method requires high-performance liquid chromatography units and tandem mass spectrometry, which are highly specialized techniques that require substantial technical and methodological considerations needed for adapting the D_3_Cr method. For perspective into the different methods included in the current study, **Figure 1** shows a pro/con list comparing the D_3_Cr dilution method in addition to other procedures like DXA and BIA, where all methods estimate skeletal muscle mass [15].

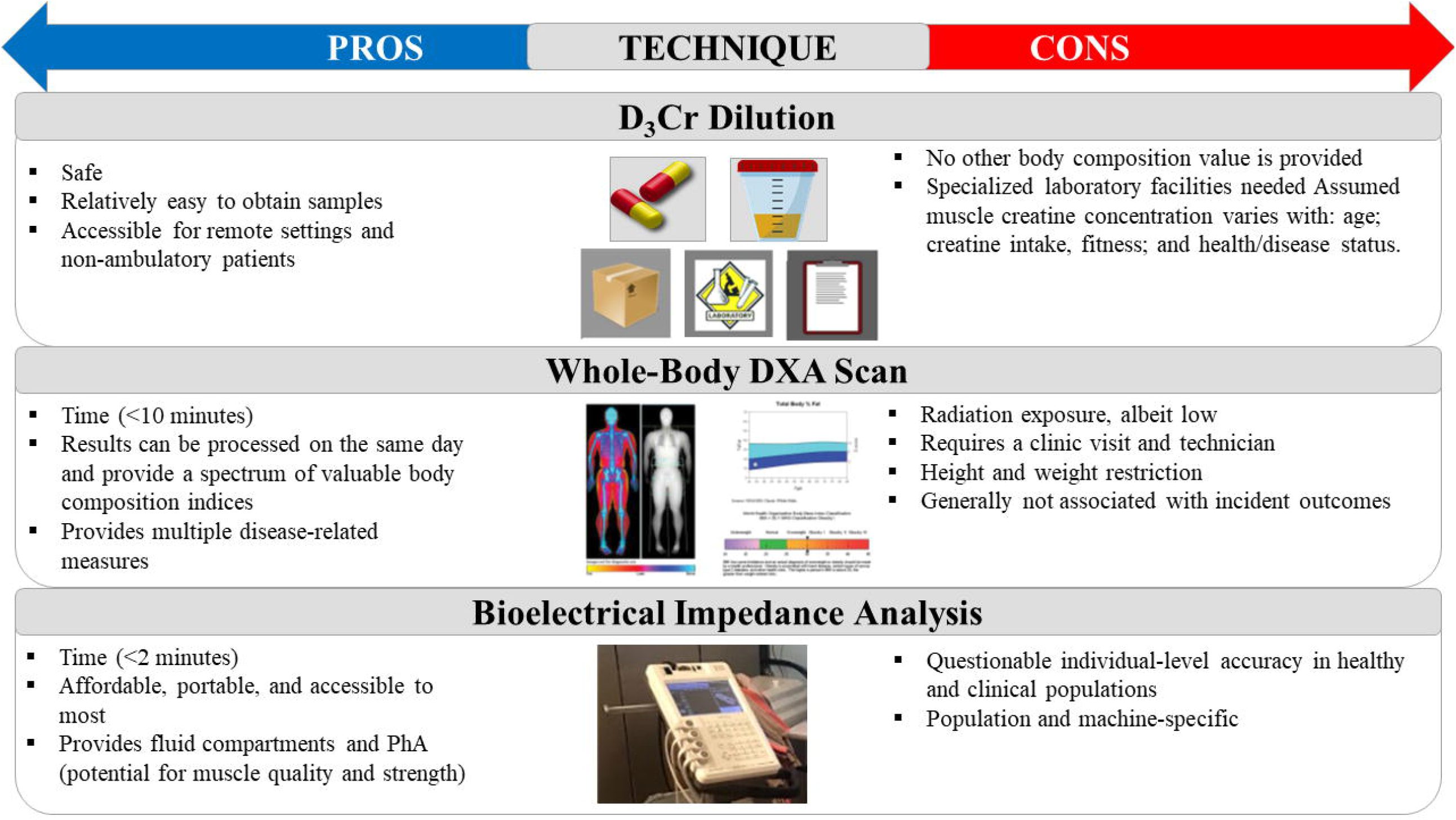

D_3_Cr has been reported to provide an accurate assessment of skeletal muscle mass with associations to MRI muscle volume in older males/females, post-menopausal females, young males (*r*=0.87) [16], and athletes (*r*=0.90) [17]. D_3_Cr muscle mass has associations with DXA ALST (*r*=0.58), grip strength (*r*=0.20), walking speed (*r*=0.29) in older males [18], and physical function in boys with Duchenne muscular dystrophy [19]. As a result, D_3_Cr shows promise for use in muscle assessment in clinical and field research [20]. Research further suggests that in all quartiles of the older adult male population, DXA had weak or no associations than D_3_Cr-derived muscle mass for serious injurious falls (*p*-trend=0.022), grip strength and with mortality and incident disability (*p*-trend=<0.001) [21], indicating possible advantages of D_3_Cr for assessments of functional status and falls/fracture prevention. D_3_Cr methods have also been explored in postmenopausal females, showing a stronger relationship in physical function using the short physical performance battery test over DXA LST [22]. The comparative advantages of D_3_Cr estimates of muscle mass and physical strength in young adults and children are unknown. No other studies have shown isokinetic strength estimates using D_3_Cr in athletes. To date, the only other studies using D_3_Cr in younger populations have found associations with total body water (*r*=0.92) and body mass (BM) (*r*=0.94) [23] in infants. On the other hand, young adult athletes are an ideal opportunistic population to study since they have a vested interest in strength and functional performance [24] and because strength can vary greatly due to the specific sport itself [25].

This study seeks to clarify how DXA LST and D_3_Cr muscle mass are related to strength and muscle in young fit adults. We compared D_3_Cr, common DXA representations of LST including total body and appendicular masses, and the leg and trunk strength. Additionally, we investigated whether BIA variables explain more variability in strength than using DXA or D_3_Cr measures alone. We hypothesized that D_3_Cr and DXA LST have similar associations with strength in young adults as previously reported in older adults.

## Methods

This cross-sectional study referred to as the *Da Kine Study* investigated a sample of collegiate and intramural student-athletes who received a battery of measures related to strength and body composition including demographics, D_3_Cr, whole-body DXA, and BIA. This study was approved by the University of Hawai’i Research Compliance and Institutional Review Board (IRB), Protocol # 2018-01102. Written consent was obtained from all participants.

### Study Population

Between April 2019 and March 2020, 80 healthy male and female collegiate and intramural athletes (≥18 years) of all BMI ranges were enrolled. Athletes were recruited if they were competing in-season or participating in an off-season strength and conditioning routine specific to their sport. All sports played are represented in **Supplemental Table 1**. Recruitment was performed by investigators visiting coaches and trainers during practice times, where participants were provided with an overview of the study, the study requirements, and expected outcomes. Participants were required to fast at least eight hours before testing and avoid extraneous exercise for 24 hours before the testing day. The athletes were instructed to not consume alcohol the day before or exercise the day of the exam day. On the testing day, participants arrived at the University of Hawaii Cancer Center, where they confirmed adherence to the pretesting protocols, voided and changed into form-fitting clothing (form-fitting shorts, a sports bra for females, and a swim cap); participants were provided an exam gown for comfort during the study. Anthropometric measures and strength tests were measured by the same technician for all measures. The primary race was reported by the participant. Height and weight measurements were made using a SECA 264 (Seca, Chino, CA, USA) measuring station. Participants were excluded from participation if they were pregnant or breastfeeding, unable to perform strength measures due to injury, had metal implants (knee, hip, shoulder, etc.), or had received surgical procedures that altered body composition (ex; liposuction or breast reduction/enhancement). Full details of the testing procedure are outlined elsewhere [26].

### D_3_Cr Creatine Dilution Method to Measure Muscle Mass

Before dosing, participants were asked to abstain from supplemented creatine (> 8 weeks), and high dietary sources of creatine (>48 hours) which was verified verbally before the start of the study. D_3_Cr is a stable isotope that is given orally in water and distributed to the muscle compartment of the body. This method follows certain assumptions and is comprehensively reviewed in a recent historical review [13]. In brief, dietary creatine is typically ingested, digested, absorbed, and transported into muscle against a gradient and is metabolized into creatinine which is excreted in the urine. D_3_Cr follows the same physiological path as dietary creatine and the enrichment of labeled creatinine in the urine is then collected to determine a creatine pool size and as a result estimate whole-body muscle mass. For this study, each subject ingested a single 60 mg dose of D_3_Cr three days before laboratory testing. Fasted second-morning void of urine was collected upon arrival at the laboratory, and aliquots were stored at 20°C. Aliquots were frozen and shipped for processing at the Department of Nutritional Science and Toxicology at the University of California, Berkeley. The methodologies utilized in the *Da Kine Study* differ slightly from previous protocols (e.g., single urine spot collection), whereas the current study collected urine in 50ml urine vials, however, these differences should not have adversely influenced data quality.

As a methodological reference, muscle mass was estimated using the D_3_Cr dilution method described in the protocol outlined in previous work [27]. The D_3_Cr method to measure muscle mass had an updated algorithm that corrected for a certain loss of creatine in the urine, theoretically making the method more accurate [27]. To account for body size differences across participants, the primary outcome of D_3_Cr muscle mass and its ratio to BM were included in the analysis, following previously published procedures [18, 21].

### Dual Energy X-Ray Absorptiometry

Whole-body DXA scans were performed using a Hologic Discovery/A system (Hologic Inc., Marlborough, MA, USA) The scans were analyzed at the University of Hawaii Cancer Center by a trained technologist using Hologic Apex version 4.5, whole-body Fan Beam, with the National Health and Nutrition Examination Survey Body Composition Analysis calibration option disabled. DXA systems were calibrated according to standard Hologic protocols and all scans were taken using standardized procedures [28]. The DXA scans provided whole-body LST (with and without head based on previous studies [9, 29, 30]), ALST, and leg LST measures. Further variables were created to normalize body composition measures by BM (LST/BM and ALST/BM), height (ALST/ht^2^), and BMI (ALST/BMI) to match variables previously defined in the literature [3, 31].

### Bioelectrical Impedance

Multifrequency BIA (MF-BIA) was performed on all participants using the InBody S10 (InBody, Cerritos, CA, USA). All scans were acquired in the supine position on the DXA scan table immediately following the DXA scan and after lying flat for ten minutes. The MF-BIA system utilizes six frequencies ranging from 1kHz -1MHz for the prediction of water compartments and body composition. Each measurement was performed as per the device manufacturer’s recommendations. Measures included 50 kHz whole-body phase angle, intracellular water, extracellular water, extracellular to intracellular water (E/I) ratio, skeletal muscle mass, and ALST. The InBody S10 does not have an athlete-specific result, and so the generic measure was provided for this analysis.

### Strength Assessments

Whole-body muscle strength assessment was performed by collecting right knee and trunk extension on an isokinetic dynamometer (Humac NORM, Computer Sports Medicine, Stoughton, MA, USA). First, the concentric and eccentric strength of the knee extensors/flexors was assessed at 60 and 180°, and subsequently, concentric and eccentric tests for the trunk were performed at 60 and 120°. Participants were positioned on the Humac testing chair with a trunk-to-thigh angle of approximately 95° and fastened into the machine as per the manufacturer’s instructions. Straps were used to stabilize each participant’s lower leg, thigh, and waist. Before testing, each participant was allowed to warm up/practice to familiarize themselves with the test. After a short rest period, they performed five isometric contractions of knee extension and flexion. After another short rest period (∼five minutes), participants performed a consecutive cycle of concentric knee extension and flexion. After completing the lower body strength test, they were asked to rest and prepare for the trunk strength test. The upper body trunk strength test consisted of two parts: five repetitions of the maximal effort of trunk flexion and extension with rest and the second part of 15 consecutive repetitions. As shown in **Figure 2** trunk flexion and extension strength were measured on the modular trunk component of the Humac.

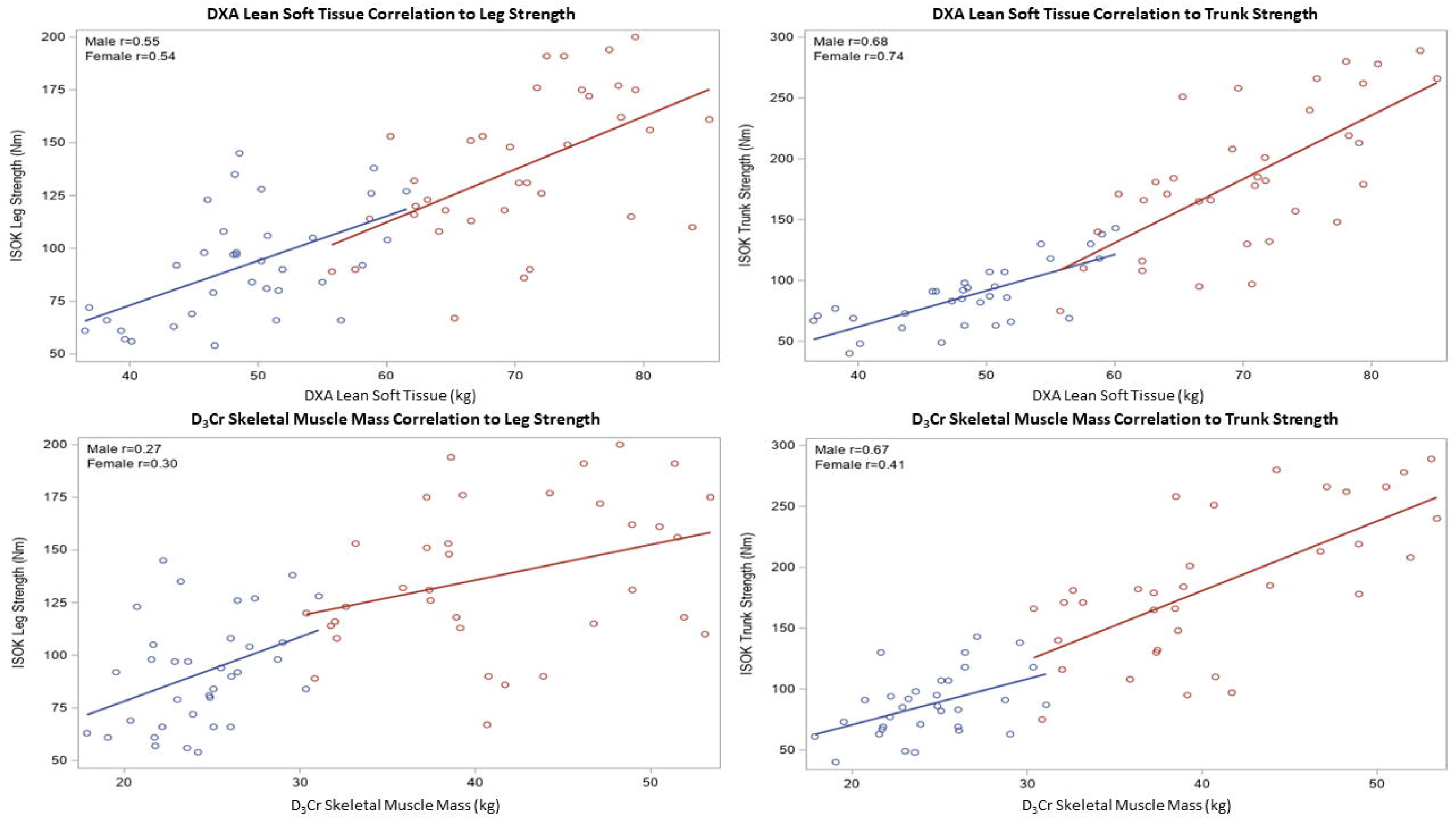

All data collection was followed by the recommended protocol of the Humac NORM manual [32]. No gravity correction was used for any tests. Before the start of the test, verbal instructions were given to push as hard and fast as possible through the full range of motion. During the test, verbal encouragement was made, but the computer screen was positioned so that participants could not see real-time feedback on their effort. Isokinetic peak torque was recorded as the maximum torque in Newton-meters (Nm) achieved during the repetitions. The primary dependent muscle strength variable was isokinetic leg and trunk extension. Isokinetic leg and trunk extension incorporate more muscles into the action, producing the largest mean Nm peak force which would be more representative of whole-body muscle mass measures.

### Statistical Analysis

Data were divided by sex because there is reason to believe the association of muscle size with muscle strength may vary, given known sex differences in strength and body composition. All measures were tested for normality of the residuals using the Shapiro–Wilks’s test and found to be normally distributed. Pearson’s product correlation coefficient was used to show determine the associations between like measures. A *p-*value *of* < 0.05 was considered significant. Generalized linear modeling was used to compute the means of the various whole-body strength tests across quartiles of body composition measures and report 95% confidence intervals and *p* trend from these models. We controlled the influence of outliers by using an a priori outliers check, defining outliers as more than 3 standard deviations from the mean value. Least squares mean (LSMEANS) were compared between groups. The base model was built using a forward stepwise selection of demographic information (age, BMI, height, and weight) and the BIA variables as a baseline comparison when all other body composition measures were added to the base model. Significance levels for entry and stay were 0.10 and 0.05 respectively. Results were reported as adjusted *R*^2^ and RMSE. Bootstrapping (n=1000) 95% confidence intervals for the *R*^2^ of each model, using the percentile method was used to report overlapping. All statistical calculations were performed using SAS 9.4 (SAS, Cary, NC, USA).

## Results

Of the 80 participants recruited, 1 was missing D_3_Cr and 2 was missing a valid trunk strength measurement. Two D_3_Cr muscle mass results were excluded from the analysis due to their non-physiological possibility. After data processing, 75 (38 females) participants were included in the final analysis, **Table 1** shows the characteristics of the study participants separated by sex. Participant recruitment with exclusions is presented in **Supplemental Figure 1**. All variables differed between sexes (*p*<0.05), and given the strong *a priori* suggestion that muscle size and strength are likely to vary between males and females justify the separate analysis by sex. All measures, including strength, were normally distributed.

**Table 1.** Descriptive Characteristics

| Variable | Males (n=37) |  |  |  | Females (n=38) |  |  |  |
| --- | --- | --- | --- | --- | --- | --- | --- | --- |
|  | Mean | SD | Min | Max | Mean | SD | Min | Max |
| Weight (kg) | 81.29 | 9.71 | 61.90 | 101.25 | 62.88 | 11.22 | 43.90 | 101.80 |
| Height (cm) | 180.31 | 10.33 | 159.30 | 203.00 | 167.35 | 8.11 | 154.70 | 185.30 |
| Age (years) | 24.27 | 4.80 | 18.00 | 36.00 | 21.86 | 4.25 | 18.00 | 35.00 |
| BMI (kg/m<sup>2</sup>) | 25.11 | 3.23 | 20.15 | 32.79 | 22.41 | 3.32 | 17.79 | 32.24 |
| Leg ISOK Ex (Nm) | 138.36 | 34.64 | 67.00 | 200.00 | 91.49 | 25.75 | 54.00 | 145.00 |
| Trunk ISOK Ex (Nm) | 273.71 | 89.89 | 137.00 | 494.00 | 165.16 | 44.41 | 72.00 | 259.00 |
| D<sub>3</sub>Cr SMM (kg) | 41.30 | 7.06 | 30.37 | 53.40 | 24.35 | 3.19 | 17.87 | 31.08 |
| D<sub>3</sub>Cr SMM/ BM (kg) | 0.51 | 0.07 | 0.38 | 0.68 | 0.39 | 0.06 | 0.26 | 0.55 |
| DXA LST (kg) | 67.35 | 7.16 | 52.90 | 81.67 | 46.38 | 6.30 | 34.82 | 59.07 |
| DXA Right Leg LST (kg) | 70.43 | 7.46 | 55.74 | 85.11 | 48.65 | 6.56 | 36.52 | 61.55 |
| DXA ALST (kg) | 12.11 | 1.49 | 9.42 | 15.07 | 8.41 | 1.20 | 5.71 | 11.20 |
| DXA ALST/ BM (kg) | 33.40 | 3.78 | 25.53 | 41.21 | 21.84 | 3.25 | 15.05 | 28.50 |
| DXA ALST/ht<sup>2</sup>(kg/m<sup>2</sup>) | 0.41 | 0.03 | 0.33 | 0.46 | 0.35 | 0.04 | 0.25 | 0.42 |
| DXA ALST/ BMI | 10.30 | 1.12 | 8.39 | 12.83 | 7.79 | 0.94 | 5.65 | 9.56 |
| BIA PhA (°) | 1.35 | 0.19 | 0.91 | 1.70 | 0.98 | 0.13 | 0.72 | 1.25 |
| BIA ICW (L) | 7.30 | 0.65 | 5.70 | 8.70 | 6.51 | 0.79 | 5.50 | 9.80 |
| BIA ECW (L) | 31.65 | 3.74 | 24.30 | 38.40 | 21.67 | 3.18 | 15.40 | 27.30 |
| BIA SMM (kg) | 39.28 | 4.88 | 29.60 | 48.00 | 26.27 | 4.14 | 18.10 | 33.60 |
| BIA E/I | 0.59 | 0.02 | 0.56 | 0.64 | 0.60 | 0.01 | 0.58 | 0.63 |

Abbreviations: ALST – appendicular lean soft tissue, BM – body mass, BMI – body mass index, LST – lean soft tissue, SMM – skeletal muscle mass, ECW – extracellular water, E/I – extracellular-to-intracellular water ratio, ICW – intracellular water, ECW – extracellular water, PhA – phase angle, SD – standard deviation, ISOK – isokinetic, ISOM – isometric, Flex – flexion, Ex – extension

**Table 2** shows the Pearson correlations and 95% confidence intervals of D_3_Cr muscle mass and selected DXA measures to whole-body muscular strength outcomes, with values significantly different from zero (*p*<0.05) bolded. Demographic information was included in the analysis as a point of reference between other body composition measures. Age, D_3_Cr/BM, ALST/BMI, phase angle, and intracellular water were not statistically correlated to any strength measure. Significant correlations were not observed for D_3_Cr muscle mass and male and female leg strength. When comparing group correlations between male leg to female leg and male trunk to female trunk, the Pearson correlation coefficient of all measures (D_3_Cr, DXA, and BIA) was generally within the same range as each other. However, when comparing leg strength to trunk measures, trunk strength was generally more highly associated with all body composition measures. There were neither discernable nor unique differences in the associations of DXA LST (with and without head) and strength measures (without head LST not shown). There was a large overlapping effect of the 95% confidence intervals per variable comparison, which implies that each variable may not have a statistically significant difference. For example, all measures of DXA LST overlapped with measures of D_3_Cr, however, DXA LST confidence intervals did not overlap with other measures like DXA ALST/ht2. Further strength analyses such as isokinetic trunk and leg flexion strength and isometric leg flexion/extension measures were performed in the *Da Kine Study* and are shown in **Supplemental Table 2** in males and **Supplemental Table 3** in females. For the whole-body variables of interest, partial correlations were performed by adjusting for height and weight which can be found in Supplemental Table 4 in males and Supplemental Table 5 in females. DXA LST, right leg LST, ALST, and ALST/BM all showed significant correlations in both sexes across all measures of isokinetic/isometric strength in both leg and trunk strength. Age, D_3_Cr/BM, and ALST/BMI were all non-significant in both sexes across all measures of isokinetic/isometric strength in both leg and trunk strength.

**Table 2.** Pearson’s Correlation (r) of D_3_Cr Muscle Mass, DXA and BIA Measures to Isokinetic Strength

| Parameter | Male |  |  |  | Female |  |  |  |
| --- | --- | --- | --- | --- | --- | --- | --- | --- |
|  | Leg ISOK Ex |  | Trunk ISOK Ex |  | Leg ISOK Ex |  | Trunk ISOK Ex |  |
|  | *r* | 95%CI | *r* | 95%CI | *r* | 95%CI | *r* | 95%CI |
| Weight (kg) | 0.15 | -0.07,0.53 | **0.40** | 0.06,0.64 | 0.36 | 0.0,0.59 | **0.46** | 0.15,0.7 |
| Height (cm) | **0.47** | 0.15,0.68 | **0.60** | 0.29,0.76 | 0.27 | 0.02,0.61 | **0.68** | 0.42,0.82 |
| Age (years) | 0.07 | -0.29,0.35 | -0.09 | -0.41,0.24 | -0.11 | -0.48,0.16 | 0.05 | -0.46,0.21 |
| BMI (kg/m<sup>2</sup>) | **0.34** | -0.11,0.51 | 0.23 | -0.13,0.5 | 0.11 | -0.12,0.51 | **0.50** | 0.16,0.71 |
| D<sub>3</sub>Cr SMM (kg) | 0.27 | 0.0,0.6 | **0.67** | 0.42,0.82 | 0.30 | 0.05,0.63 | **0.41** | 0.13,0.69 |
| D<sub>3</sub>Cr SMM /BM (kg) | 0.11 | -0.28,0.38 | -0.28 | -0.05,0.57 | 0.08 | -0.42,0.23 | 0.38 | -0.59,0.04 |
| DXA LST (kg) | **0.55** | 0.26,0.74 | **0.68** | 0.44,0.82 | **0.54** | 0.25,0.74 | **0.74** | 0.51,0.86 |
| DXA Right Leg LST (kg) | **0.55** | 0.14,0.68 | **0.69** | 0.3,0.76 | **0.54** | 0.33,0.77 | **0.73** | 0.49,0.85 |
| DXA ALST (kg) | **0.46** | 0.23,0.72 | **0.59** | 0.37,0.79 | **0.55** | 0.28,0.75 | **0.69** | 0.53,0.86 |
| DXA ALST / BM (kg) | **0.53** | -0.04,0.56 | **0.64** | -0.06,0.55 | **0.50** | 0.11,0.66 | **0.72** | 0.21,0.73 |
| DXA ALST /ht<sup>2</sup>(kg/m<sup>2</sup>) | **0.43** | -0.27,0.38 | 0.28 | -0.31,0.35 | 0.34 | -0.15,0.49 | **0.51** | -0.36,0.32 |
| DXA ALST/ BMI (kg/kg/m<sup>2</sup>) | 0.06 | -0.35,0.31 | 0.01 | -0.27,0.38 | 0.21 | -0.16,0.51 | -0.09 | -0.24,0.47 |
| BIA PhA (°) | 0.24 | 0.09,0.65 | 0.32 | 0.39,0.8 | 0.28 | 0.04,0.64 | 0.22 | 0.29,0.79 |
| BIA ICW (L) | 0.03 | 0.13,0.68 | 0.06 | 0.35,0.78 | 0.23 | 0.06,0.65 | 0.13 | 0.29,0.78 |
| BIA ECW (L) | **0.39** | -0.15,0.48 | **0.65** | -0.41,0.24 | 0.35 | -0.31,0.38 | **0.60** | -0.48,0.23 |
| BIA SMM (kg) | **0.40** | 0.1,0.66 | **0.65** | 0.39,0.8 | 0.35 | 0.04,0.64 | **0.59** | 0.28,0.78 |
| BIA E/I | 0.30 | 0.09,0.65 | **0.55** | 0.39,0.8 | **0.48** | 0.04,0.64 | **0.62** | 0.29,0.79 |

Abbreviations: ALST – appendicular lean soft tissue, BM – body mass, BMI – body mass index, LST – lean soft tissue, SMM – skeletal muscle mass, ECW – extracellular water, E/I – extracellular-to-intracellular water ratio, ICW – intracellular water, ECW – extracellular water, PhA – phase angle, SD – standard deviation, ISOK – isokinetic, ISOM – isometric, Flex – flexion, Ex – extension
Bolded values indicate statistical significance (p < 0.05)

**Table 3** shows the results of the model performance of the base model and the additional body composition measures (DXA and D_3_Cr). Stepwise forward regression selected only height and weight as the candidate variables to form the base mode, where all other variables, including BIA variables, were not included. DXA LST had the highest leg (*R*^*2*^=0.36, 0.37) and trunk (*R*^*2*^=0.53, 0.61) strength in both males and females. However, DXA right leg LST had the highest *R*^*2*^ for females’ leg strength (*R*^2^=0.40). Trunk strength was more highly associated with body composition measures than leg strength in both sexes and all measurement techniques. One or more of the DXA LST measures (total, leg, and ALST) was consistently more highly associated with leg and trunk strengths for both sexes than BIA and D_3_Cr measures. Significant increases in model *R*^2^ were not observed due to all variables having large overlapping effects of the 95% *R*^2^ confidence intervals, including the base model.

**Table 3.** Regression Results of D_3_Cr Muscle Mass to DXA Measures to Strength

| Parameter | Male |  |  |  |  |  | Female |  |  |  |  |  |
| --- | --- | --- | --- | --- | --- | --- | --- | --- | --- | --- | --- | --- |
|  | Leg ISOK Ex |  |  | Trunk ISOK Ex |  |  | Leg ISOK Ex |  |  | Trunk ISOK Ex |  |  |
|  | R<sup>2</sup> | 95% CI | RMSE | R<sup>2</sup> | 95% CI | RMSE | R<sup>2</sup> | 95% CI | RMSE | R<sup>2</sup> | 95% CI | RMSE |
| Base Model (Model 0) | 0.25 | 0.05-0.54 | 31.3 | 0.38 | 0.13-0.66 | 49.1 | 0.18 | 0.02-0.49 | 24.4 | 0.49 | 0.2-0.73 | 19.7 |
| Model0+D<sub>3</sub>Cr SMM | 0.27 | 0.07-0.56 | 33.5 | 0.49 | 0.27-0.71 | 44.0 | 0.26 | 0.15-0.64 | 22.0 | 0.54 | 0.26-0.78 | 19.4 |
| Model0+DXA LST | 0.36 | 0.15-0.62 | 29.3 | 0.53 | 0.3-0.74 | 45.2 | 0.37 | 0.05-0.57 | 24.2 | 0.61 | 0.36-0.8 | 17.4 |
| Model0+DXA Right Leg LST | 0.29 | 0.08-0.57 | 31.3 | 0.42 | 0.18-0.66 | 48.8 | 0.40 | 0.18-0.64 | 20.9 | 0.57 | 0.3-0.78 | 18.0 |
| Model0+DXA ALST | 0.33 | 0.12-0.6 | 30.0 | 0.44 | 0.21-0.68 | 46.7 | 0.36 | 0.13-0.63 | 21.6 | 0.60 | 0.34-0.8 | 17.8 |
| Model0+BIA SMM | 0.29 | 0.08-0.58 | 31.4 | 0.47 | 0.23-0.71 | 46.0 | 0.24 | 0.04-0.6 | 31.4 | 0.52 | 0.22-0.77 | 46.0 |

Abbreviations: ALST – appendicular lean soft tissue, RL – Right Leg, SMM – skeletal muscle mass, LST – lean soft tissue, SMM – skeletal muscle mass, ICW – intracellular water. ISOK – isokinetic, ISOM – isometric, Flex – flexion, Ex – extension
Base Model included height and weight variables. RMSE units are in Nm. All models are reporting adjusted R<sup>2</sup>

D_3_Cr muscle mass consistently performed lower than DXA variables, in line with Table 2 which showed non-significant Pearson’s correlations with D_3_Cr muscle mass in both male and female leg strength. All of the BIA variables performed similarly to the DXA variables but were not superior to DXA LST. Visualization of the variance in the data is described in **Figure 3** as a scatterplot of DXA LST and D3Cr muscle mass to leg and trunk strength. Large variance is seen within the males over the females in leg and trunk strength, with DXA LST showing less variance over D_3_Cr muscle mass.

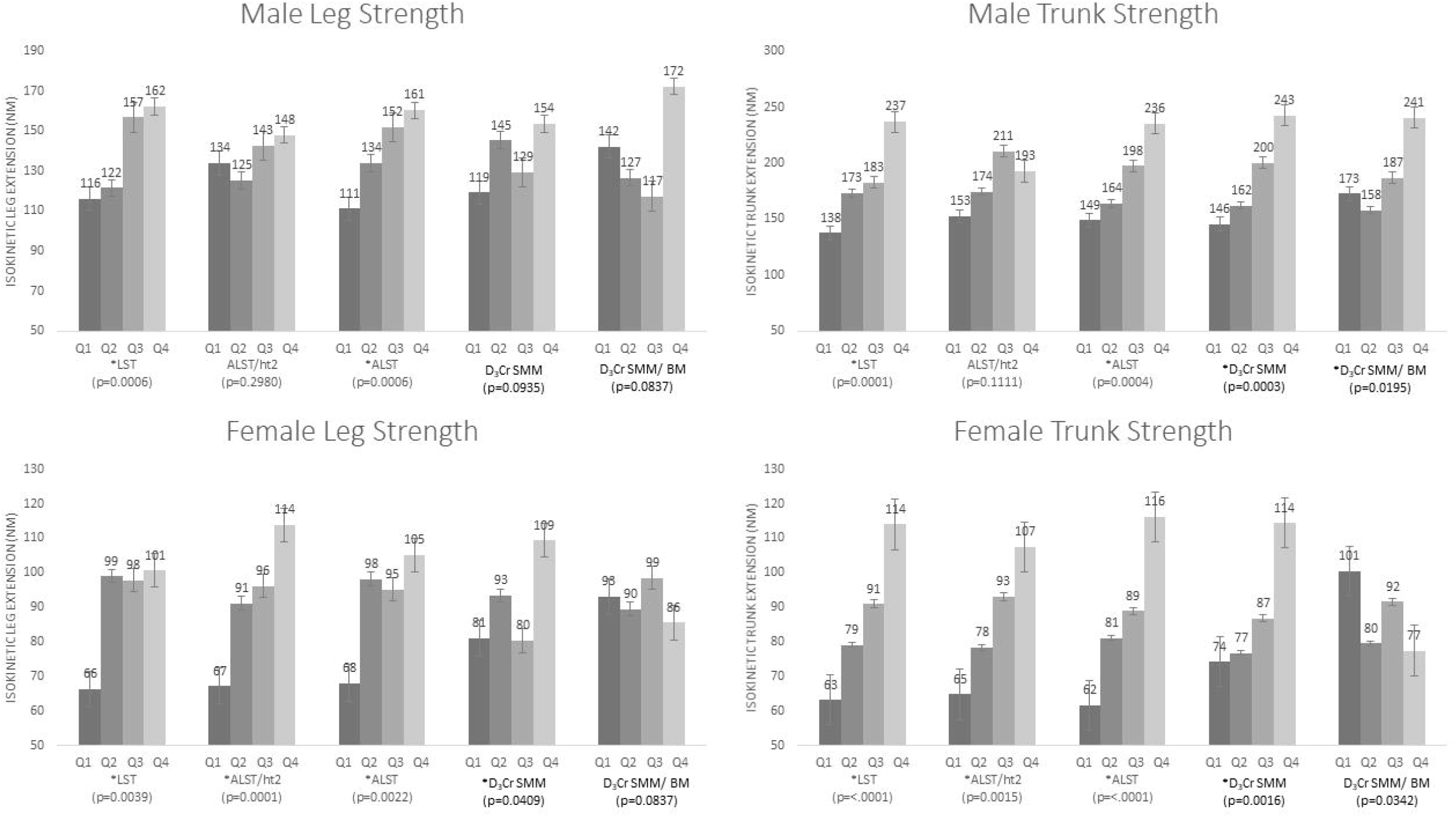

**Figure 4** shows the quartiles of body composition measures related to strength. Males showed a significant *p* trend for all DXA variables except for ALST/ht^2^ in both strength measures. Only DXA LST and ALST showed a significant *p* trend throughout measures of strength in both males and females, suggesting the importance of using LST measures, as opposed to normalized LST measures. In males, D_3_Cr showed a significant *p* trend for trunk strength in both D_3_Cr/BM and D_3_Cr alone but not for leg strength. In females, DXA and D_3_Cr variables had a significant *p* trend in both strength measures, except for D_3_Cr/BM.

## Discussion

The purpose of this study was to explore the use of clinical and laboratory-based whole and regional body composition measures associations with muscle strength in young fit adults and contrast these results with published findings in older adults. Our study showed that DXA LST was more highly associated with both leg and trunk strength than D_3_Cr or other candidate measures, with or without using adjustment for demographics, or BIA variables. The only exception was a higher association of female DXA right leg LST with leg strength versus whole-body LST. Further, all body composition predictors had a higher association with trunk strength than leg strength for both males and females. Lastly, ALST/ht^2^, commonly used in definitions of sarcopenia [3], was not as highly associated with strength in general as DXA right leg LST and total LST (Table 2), underscoring the limitation of using this approach of normalizing body composition measures to height in the associations of absolute strength. Raymond-Pope found similar findings to our study in that right leg LST from DXA correlated with right leg isokinetic extension with an *r*=0.80 in female college athletes [33]. Further, Bourgeois found comparable findings to ours who used right leg LST associations to leg strength with an *R*^2^=0.57 in an adult population [8].

There are potential implications of comparing upper and lower body relationships within populations such as athletes and older adults, such that lower body muscle strength is typically wasted faster in older adults [34]. Grip strength is routinely used in older populations as a determinant of poor outcomes and as part of sarcopenia definitions; however, calf circumference and lower extremity strength may be more important for mobility outcomes [35]. In the current study, the sports that were collected could be less related to lower-body strength and more related to upper-body strength, or vice versa. In other words, different sports require different body adaptations; so, the sport may have less of a demand on an individual’s upper body or lower body (ex; soccer vs. baseball). We found that trunk strength was more highly associated with using body composition measures over leg strength in both sexes.

The combination of DXA and BIA has previously been shown to increase the association of DXA measures and muscle strength in older adults by using variables such as phase angle, intracellular and extracellular water, or the ratio of the two [9, 11]. We found that phase angle showed no association with all measures of strength, whereas the water compartments had significant correlations to muscle strength, with the extracellular fluid being the highest. Furthermore, none of the BIA variables were selected in the forward stepwise regression modeling to improve either DXA and D_3_Cr associations to muscle strength. These results point to potential physiological differences between athletes and the literature for older adults, where BIA variables such as phase angle may serve as a better tool to reflect muscle strength. The relationship between muscle mass and strength remains unclear despite a positive linear association between mass and strength [36]. This may be the result of the type of strength being assessed [37], different types of sport-specific training, and consequently muscle fiber types and their contribution to strength versus power [38], the effect of increased fat deposition in muscle, associated with greater fat oxidation in athletes [39], or the effect of LST hydration [40]. However, these findings support that LST reflects muscle strength in this population.

Results of the D_3_Cr performance compared to DXA for associations with strength and functional measures have been mixed in older adult populations most likely due to several factors. The studies available that compare D_3_Cr, DXA, and strength measures generally show a higher association for DXA measures than for D_3_Cr. One study on older adults performed knee isokinetic strength tests of one repetition maximum and maximum voluntary contraction (MVC) and found that D_3_Cr muscle mass showed a slightly higher association over measures from DXA LST and ALST (MVC: *R*^*2*^=0.70, 0.65, 0.63 respectively, [41]). Grip strength does not test major muscle groups that directly resist gravity but is a more accessible measure and commonly utilized in sarcopenia studies instead of trunk and leg strength. For example, the males in the MrOS study underwent D_3_Cr, whole-body DXA, and grip strength where it was observed that the trend of increasing DXA ALST/ht^2^ was more significantly associated with grip strength than for D_3_Cr muscle mass/BM (p<0.001 versus 0.002 respectively) [21]. Buehring et al. [30] found that in males and females over 70 years, DXA LST had a higher *R*^*2*^ than D_3_Cr in maximum hand grip strength (DXA LST *R*^*2*^=0.40, D_3_Cr *R*^*2*^=0.12).

However, studies that compared D_3_Cr muscle mass, DXA measures, and functional tests often show a stronger association between D_3_Cr and function than DXA [42-44]. The males in the MrOS study underwent functional measures including walking speed, 6-minute walk, chair-to-sit tests, and a jump test for lower extremity power, all commonly used measures of functional limitations in older adult populations [21]. The D_3_Cr measures showed a significant trend to lower extremity power (watts/kg) and short physical performance battery score while ALST/ht^2^ showed no association. Other studies showed a similar superior association of D_3_Cr to functional tests versus DXA measures [42-44]. In another study, D_3_Cr muscle mass/BM was shown to be more highly associated with habitual walking speed (*p*=0.038) than DXA LST (*p*=0.513), ALST (*p*=0.573), ALST/h^2^ (*p*=0.604) or ALST/BM (*p*=0.069) in an older adult population [18]. One contrasting study is Buehring. [30] who found that DXA LST had a higher *R*^*2*^ than D_3_Cr for maximum jump power (DXA LST *R*^*2*^=0.40, D_3_Cr *R*^*2*^=0.34) [30] in older males and females. We have no comparable functional measures in our study.

Many of the studies referred to above report results for either both sexes combined (e.g. Buehring, Raymond-Pope, and Bourgeois) or just for one sex, such as the males in the MrOS study. Considerable sex differences related to body size are relevant when predicting muscle strength with body composition. In our study, right leg LST was associated with leg strength with an *R*^*2*^=0.36 and 0.37 for males and females respectively. Non-significant relationships, or those with relatively low correlation, may become significant when subgroups like sex are combined [45]. The combined association for males and females for right leg LST and leg strength was *R*^*2*^= 0.53, virtually identical to Bourgeois. We recommend reporting sex-specific muscle strength associations to avoid this type of correlation inflation and to not generalize results seen in males for females.

The majority of D_3_Cr research to evaluate associations to physical performance to date has been conducted in older adults and by using fundamentally different performance metrics in comparison to the current study, such as the six-minute walk, lower extremity power, short physical performance battery score, or hand grip strength tests. Further, the units of lean and muscle measure varied greatly across studies. In our study, we found that D_3_Cr performed better than D_3_Cr/BM and that DXA ALST and LST performed better than ALST/ht^2^ for strength associations. We also found that the normalized fractions of LST/BM and D_3_Cr muscle mass/BM did not show statistical significance to the leg and trunk strength measures while other studies found significant associations to grip strength and function.

The MrOs studies often normalized D_3_Cr by BM, and ALST was normalized by height squared, fundamentally different normalizations. The use of different normalizations for what are similar measures of muscle is questionable. Differences in units and normalizations may partly explain the mixed results between studies. Based on the similarity of our findings in young adults to previous studies in older adults where the same measures are presented, we predict that our findings in young adults will also hold for older adults if similar comparisons were made.

Our study collected body composition and strength measures from a racially-diverse group of collegiate athletes who participate in multiple different sports. To our knowledge, this is the first investigation evaluating the relationships between D_3_Cr measures of muscle mass and DXA for their ability to determine muscle strength in a young athletic population. Another strength of our study is that both males and females were separated in this analysis to more accurately report expected correlations by sex. Unlike DXA and BIA, D_3_Cr collection in athletes comes with strict protocol adherence that can corrupt the data. For example, creatine supplementation is very common in sports and even high dietary creatine sources. Thereby, thorough investigations on dietary creatine and supplementation are warranted in this demographic [17]. Limitations to our study include the following. A comprehensive dietary record was not collected in these participants and therefore we were unable to assure creatine consumption of the participants. Furthermore, two male participants were excluded due to a non-physiologically possible determination of muscle mass by D_3_Cr, which was inferred by the processing laboratory that these participants may have been supplementing with dietary creatine. This underscores the importance of following protocol when collecting D_3_Cr data from athletes to avoid potential data corruption. Further limitations include hand-held dynamometry and functional measures commonly used to assess older adults were not collected in our study. Although we feel that measurement of the thigh and abdominal/back muscles are more functionally relevant than grip strength, we were unable to directly compare our results too much of the sarcopenic literature. It is unclear if the results presented in athletes would be generalizable to older adults, and suggest further investigation. Finally, an inability to test the models due to the small sample size which would likely display an underpowered approach for the p-trend and potentially other results.

## Conclusion

From this investigation, we conclude that DXA standard body composition output variables, specifically LST, provide a higher relationship with the strength of the major muscle groups of the abdomen and legs compared to D_3_Cr muscle mass, BIA, and demographic variables in both male and female collegiate athletes; however, these associations were not statistically different from one another. Furthermore, we found that whole body values from all methods that were not adjusted for demographic information or BIA variables showed higher associations to muscle strength in both males and females. Future research in athletes should examine the effects of changes in muscle mass due to training, weight loss, and/or gain on LST and functional measures.

## Data Availability

All data produced in the present study are available upon reasonable request to the authors

https://shepherdresearchlab.org/

## Acknowledgment

We gratefully acknowledge En Liu and Nisa Kelly for subject recruitment and implementation of the study protocol. We thank the lab of Dr. William Evans and Dr. Mahalakshmi Shankaran for processing our samples and providing the D_3_Cr muscle mass results. The authors of this manuscript certify that they comply with the ethical guidelines for authorship and publishing in the Journal of Cachexia, Sarcopenia and Muscle [46].

## Ethical Standards

All authors certify that they comply with the Ethical guidelines for authorship and publishing in the Journal of Cachexia, Sarcopenia and Muscle.

## Author Contributions

**John Shepherd and Thomas Kelly:** Conceptualization, Funding acquisition **Devon Cataldi Brandon Quon and Lambert Leong:** Data curation, Formal analysis **John A Shepherd**: Investigation, Validation, Visualizations **John A Shepherd**: Methodology, Project administration, Resources, Software, Supervision All Authors: Roles/Writing - Original draft, review & editing. **Jonathan Bennett, Carla Prado**, William Evans and Steven Heymsfield: data analysis/interpretation. All authors provided a critical review of the manuscript.

## References

1. Cruz-Jentoft, A.J. and A.A. Sayer, Sarcopenia. The Lancet, 2019. 393(10191): p. 2636–2646.

2. Cruz-Jentoft, A.J., et al., Prevalence of and interventions for sarcopenia in ageing adults: a systematic review. Report of the International Sarcopenia Initiative (EWGSOP and IWGS). Age and ageing, 2014. 43(6): p. 748–759.

3. Cruz-Jentoft, A.J., et al., Sarcopenia: revised European consensus on definition and diagnosis. Age and ageing, 2019. 48(1): p. 16–31.

4. Guglielmi, G., et al., The role of DXA in sarcopenia. Aging clinical and experimental research, 2016. 28(6): p. 1047–1060.

5. Glüer, C.-C., et al., Accurate assessment of precision errors: how to measure the reproducibility of bone densitometry techniques. Osteoporosis international, 1995. 5(4): p. 262–270.

6. Kim, J., et al., Total-body skeletal muscle mass: estimation by a new dual-energy X-ray absorptiometry method. The American journal of clinical nutrition, 2002. 76(2): p. 378–383.

7. Hind, K., et al., Interpretation of dual-energy X-ray Absorptiometry-Derived body composition change in athletes: a review and recommendations for best practice. Journal of Clinical Densitometry, 2018. 21(3): p. 429–443.

8. Bourgeois, B., et al., Improved strength prediction combining clinically available measures of skeletal muscle mass and quality. Journal of cachexia, sarcopenia and muscle, 2019. 10(1): p. 84–94.

9. Kuchnia, A.J., et al., Combination of DXA and BIS body composition measurements is highly correlated with physical function—an approach to improve muscle mass assessment. Archives of Osteoporosis, 2018. 13(1): p. 1–9.

10. Crawford, R.J., et al., Manually defining regions of interest when quantifying paravertebral muscles fatty infiltration from axial magnetic resonance imaging: a proposed method for the lumbar spine with anatomical cross-reference. BMC musculoskeletal disorders, 2017. 18(1): p. 1–11.

11. Rush, B., et al., Combination of DXA and BIS predicts jump power better than traditional measures of sarcopenia. JBMR plus, 2021. 5(8): p. e10527.

12. Sawka, M.N., et al., Blood volume: importance and adaptations to exercise training, environmental stresses and trauma sickness. 2000.

13. McCarthy, C., et al., D3-creatine dilution for skeletal muscle mass measurement: historical development and current status. Journal of Cachexia, Sarcopenia and Muscle, 2022.

14. Prado, C.M. and S. von Haehling, D 3-Creatine dilution for body composition assessment: A direct take on the matter. Journal of Cachexia, Sarcopenia and Muscle, 2022.

15. McCarthy, C., et al., D3-creatine dilution for skeletal muscle mass measurement: historical development and current status. Journal of Cachexia, Sarcopenia and Muscle, 2022. 13(6): p. 2595–2607.

16. Evans, W.J., et al., D(3) -Creatine dilution and the importance of accuracy in the assessment of skeletal muscle mass. J Cachexia Sarcopenia Muscle, 2019. 10(1): p. 14–21.

17. Morris-Paterson, T.E., et al., Total body skeletal muscle mass estimated by magnetic resonance imaging and creatine (methyl-d3) dilution in athletes. Scand J Med Sci Sports, 2020. 30(3): p. 421–428.

18. Duchowny, K.A., et al., Association of change in muscle mass assessed by D3-creatine dilution with changes in grip strength and walking speed. Journal of cachexia, sarcopenia and muscle, 2020. 11(1): p. 55–61.

19. Evans, W.J., et al., Profoundly lower muscle mass and rate of contractile protein synthesis in boys with Duchenne muscular dystrophy. J Physiol, 2021.

20. Clark, R.V., et al., Total body skeletal muscle mass: estimation by creatine (methyl-d3) dilution in humans. Journal of Applied Physiology, 2014. 116(12): p. 1605–1613.

21. Cawthon, P.M., et al., Strong relation between muscle mass determined by D3-creatine dilution, physical performance, and incidence of falls and mobility limitations in a prospective cohort of older men. The Journals of Gerontology: Series A, 2019. 74(6): p. 844–852.

22. Zhu, K., et al., The Association of Muscle Mass Measured by D3-Creatine Dilution Method With Dual-Energy X-Ray Absorptiometry and Physical Function in Postmenopausal Women. J Gerontol A Biol Sci Med Sci, 2021. 76(9): p. 1591–1599.

23. Evans, W.J., et al., D3-creatine dilution for the noninvasive measurement of skeletal muscle mass in premature infants. Pediatric Research, 2021. 89(6): p. 1508–1514.

24. Schweighofer, N., et al., DXA-Derived indices in the characterisation of sarcopenia. Nutrients, 2021. 14(1): p. 186.

25. Andersen, E., R.G. Lockie, and J.J. Dawes, Relationship of absolute and relative lower-body strength to predictors of athletic performance in collegiate women soccer players. Sports, 2018. 6(4): p. 106.

26. Cataldi, D., et al., Agreement and Precision of Deuterium Dilution for Total Body Water and Multicompartment Body Composition Assessment in Collegiate Athletes. The Journal of Nutrition, 2022.

27. Shankaran, M., et al., Dilution of oral D3-Creatine to measure creatine pool size and estimate skeletal muscle mass: development of a correction algorithm. Journal of cachexia, sarcopenia and muscle, 2018. 9(3): p. 540–546.

28. Lu, Y., et al., Dual X-ray absorptiometry quality control: Comparison of visual examination and process-control charts. Journal of Bone and Mineral Research, 1996. 11(5): p. 626–637.

29. Jones, W., et al., Precision of the GE Lunar total body-less head scan for the measurement of three-compartment body composition in athletes. Journal of Clinical Densitometry, 2022.

30. Buehring, B., et al., Comparison of muscle/lean mass measurement methods: correlation with functional and biochemical testing. Osteoporosis International, 2018. 29(3): p. 675–683.

31. Heymsfield, S.B., et al., Scaling of body composition to height: relevance to height-normalized indexes. The American journal of clinical nutrition, 2011. 93(4): p. 736–740.

32. Humac, C., NormTM Testing & Rehabilitation System User Guide Model 770. Stoughton, MA, 2003.

33. Raymond-Pope, C.J., et al., Association of compartmental leg lean mass measured by dual X-ray absorptiometry with force production. The Journal of Strength & Conditioning Research, 2020. 34(6): p. 1690–1699.

34. Serra-Rexach, J.A., et al., Short-term, light-to moderate-intensity exercise training improves leg muscle strength in the oldest old: a randomized controlled trial. Journal of the American Geriatrics Society, 2011. 59(4): p. 594–602.

35. Fragala, M.S., et al., Strength and function response to clinical interventions of older women categorized by weakness and low lean mass using classifications from the Foundation for the National Institute of Health sarcopenia project. Journals of Gerontology Series A: Biomedical Sciences and Medical Sciences, 2015. 70(2): p. 202–209.

36. Akagi, R., et al., Muscle volume compared to cross-sectional area is more appropriate for evaluating muscle strength in young and elderly individuals. Age and ageing, 2009. 38(5): p. 564–569.

37. Evangelidis, P.E., et al., Strength and size relationships of the quadriceps and hamstrings with special reference to reciprocal muscle balance. European journal of applied physiology, 2016. 116(3): p. 593–600.

38. Hahn, T., A. Foldspang, and T. Ingemann-Hansen, Dynamic strength of the quadriceps muscle and sports activity. British journal of sports medicine, 1999. 33(2): p. 117–120.

39. Amati, F., et al., Skeletal muscle triglycerides, diacylglycerols, and ceramides in insulin resistance: another paradox in endurance-trained athletes? Diabetes, 2011. 60(10): p. 2588–2597.

40. St-Onge, M.-P., et al., Dual-energy X-ray absorptiometry lean soft tissue hydration: independent contributions of intra-and extracellular water. American Journal of Physiology-Endocrinology and Metabolism, 2004. 287(5): p. E842–E847.

41. Cegielski, J., et al., The Combined Oral Stable Isotope Assessment of Muscle (COSIAM) reveals D-3 creatine derived muscle mass as a standout cross-sectional biomarker of muscle physiology vitality in older age. GeroScience, 2022: p. 1–10.

42. Orwoll, E., et al., CT muscle density, D3Cr muscle mass and body fat associations with physical performance, mobility outcomes and mortality risk in older men. J Gerontol A Biol Sci Med Sci, 2021.

43. Zanker, J., et al., Factor analysis to determine relative contributions of strength, physical performance, body composition and muscle mass to disability and mobility disability outcomes in older men. Experimental Gerontology, 2022. 161: p. 111714.

44. Zanker, J., et al., Walking speed and muscle mass estimated by the D3-Creatine dilution method are important components of sarcopenia associated with incident mobility disability in older men: A classification and regression tree analysis. Journal of the American Medical Directors Association, 2020. 21(12): p. 1997–2002. e1.

45. Makin, T.R. and J.-J.O. de Xivry, Ten common statistical mistakes to watch out for when writing or reviewing a manuscript. Elife, 2019. 8.

46. von Haehling, S., et al., Ethical guidelines for publishing in the Journal of Cachexia, Sarcopenia and Muscle: update 2017. 2017, Wiley Online Library. p. 1081–1083.

