## Supplementary figures and images for "Association of Muscle Strength to Body Composition Measures using DXA, D_3_Cr, and BIA in Collegiate Athletes"

### Sup. Fi. 1

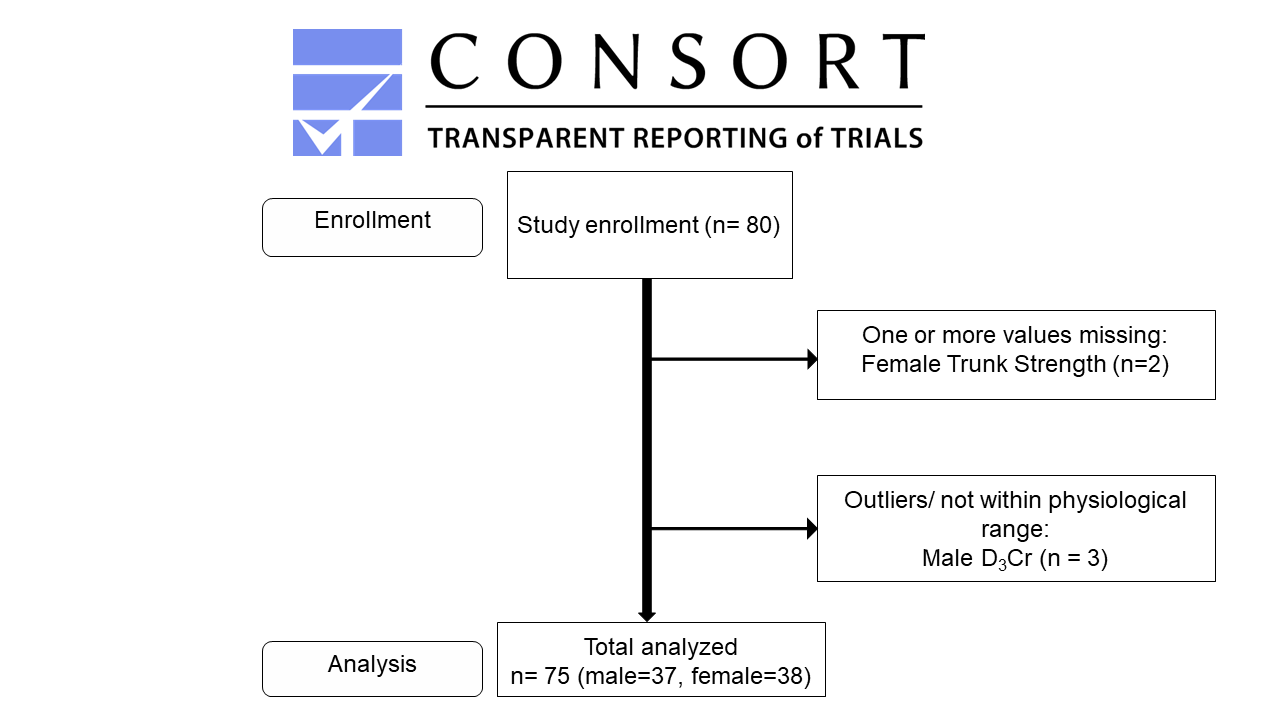

### Sup. Table 1

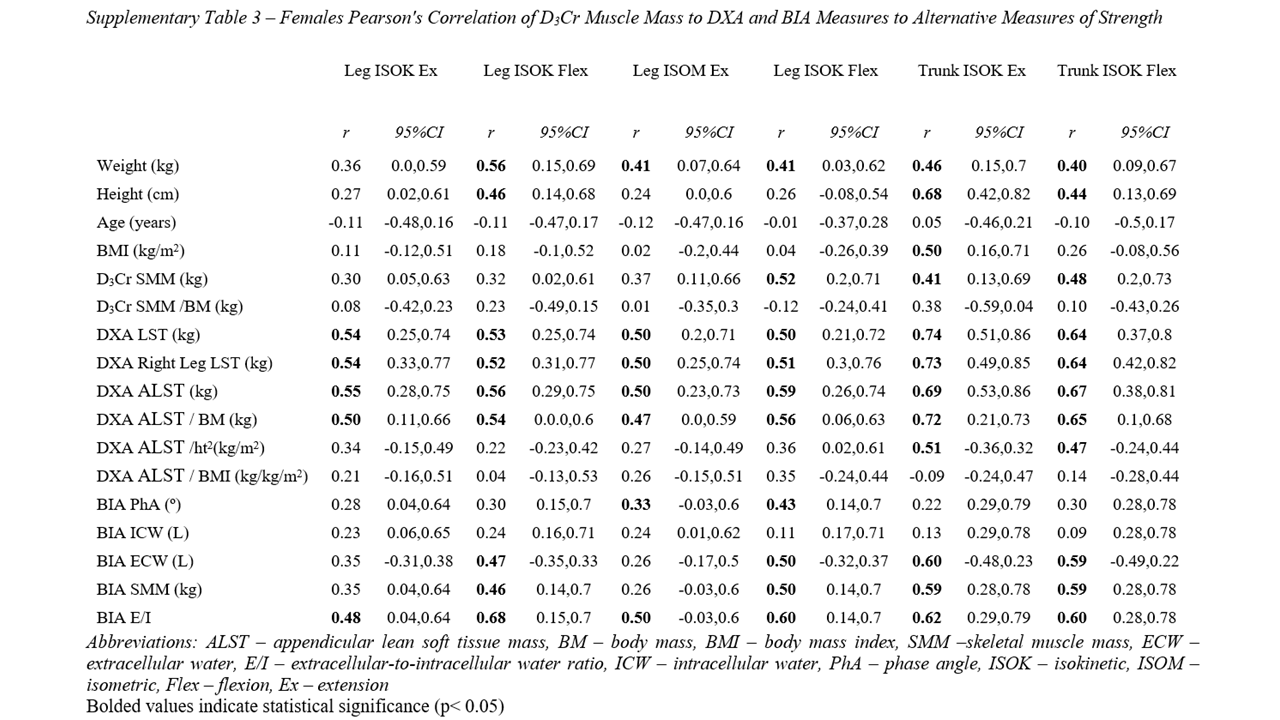

### Sup. Table 2

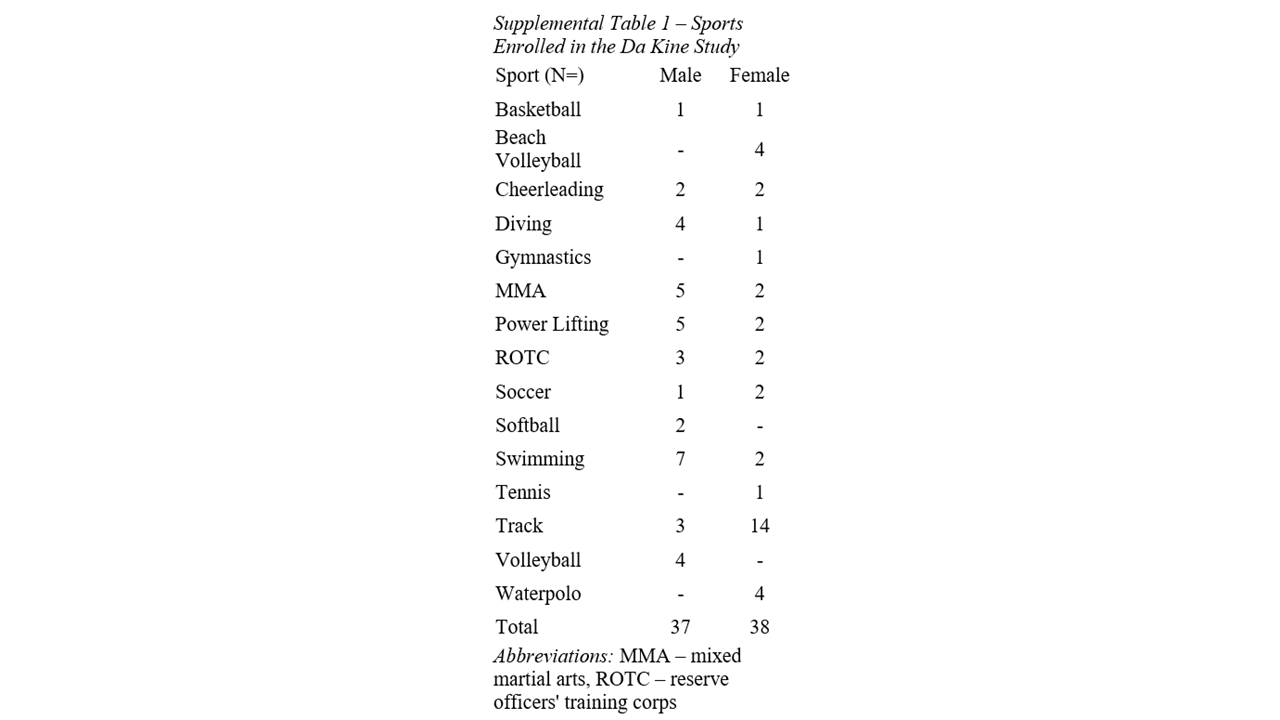

### Sup. Table 3

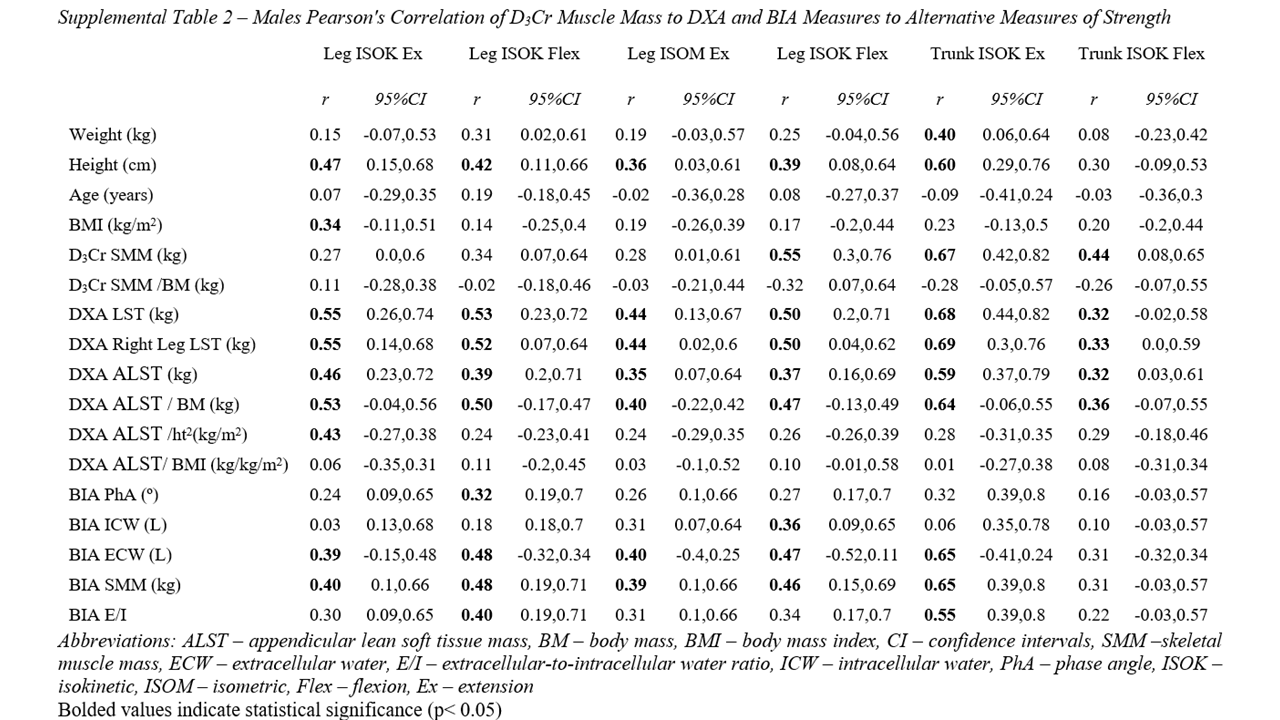
